# Self-rated smell ability enables highly specific predictors of COVID-19 status: a case control study in Israel

**DOI:** 10.1101/2020.07.30.20164327

**Authors:** Noam Karni, Hadar Klein, Kim Asseo, Yuval Benjamini, Sarah Israel, Musa Nimri, Keren Olstein, Ran Nir-Paz, Alon Hershko, Mordechai Muszkat, Masha Y. Niv

**Affiliations:** Department of Medicine, Hadassah University Hospital, Mt. Scopus Campus, Jerusalem, Israel; The Institute of Biochemistry, Food and Nutrition, The Hebrew University, Rehovot, Israel; Department of Statistics, The Hebrew University, Mt. Scopus Campus, Jerusalem, Israel; Department of Clinical Microbiology and Infectious Diseases Hadassah-Hebrew University medical center

**Author notes:** CORRESPONDING AUTHORS: Prof, Masha Y Niv, PhD, Chemical Senses and Molecular Recognition Lab, Robert H. Smith Faculty of Agriculture, Food and Environment, Rehovot, Israel, The Hebrew University of Jerusalem, Israel, Prof. Mordechai Muszkat, MD, Head, Department of Internal Medicine Hadassah University Hospital, Mt. Scopus Jerusalem, Israel. equal contributions (Noam Karni, Hadar Klein).

**Keywords:** COVID-19, smell loss, taste loss, symptoms, diagnosis, prediction, olfactory dysfunction, gustatory dysfunction, classifier, screening, rating

## Abstract

**Background:** Clinical diagnosis of COVID-19 poses an enormous challenge to early detection and prevention of COVID-19, which is of crucial importance for pandemic containment. Cases of COVID-19 may be hard to distinguish clinically from other acute viral diseases, resulting in an overwhelming load of laboratory screening. Sudden onset of taste and smell loss emerge as hallmark of COVID-19. The optimal ways for including these symptoms in the screening of suspected COVID-19 patients should now be established.

**Methods:** We performed a case-control study on patients that were PCR-tested for COVID-19 (112 positive and 112 negative participants), recruited during the first wave (March 2020 – May 2020) of COVID-19 pandemic in Israel. Patients were interviewed by phone regarding their symptoms and medical history and were asked to rate their olfactory and gustatory ability before and during their illness on a 1-10 scale. Prevalence and degrees of symptoms were calculated, and odds ratios were estimated. Symptoms-based logistic-regression classifiers were constructed and evaluated on a hold-out set.

**Results:** Changes in smell and taste occurred in 68% (95% CI 60%-76%) and 72% (64%-80%), of positive patients, with 24 (11-53 range) and 12 (6-23) respective odds ratios. The ability to smell was decreased by 0.5±1.5 in negatives, and by 4.5±3.6 in positives, and to taste by 0.4±1.5 and 4.9±3.8, respectively (mean ± SD). A penalized logistic regression classifier based on 5 symptoms (degree of smell change, muscle ache, lack of appetite, fever, and a negatively contributing sore throat), has 66% sensitivity, 97% specificity and an area under the ROC curve of 0.83 (AUC) on a hold-out set. A classifier based on degree of smell change only is almost as good, with 66% sensitivity, 97% specificity and 0.81 AUC. Under the assumption of 8% positives among those tested, the predictive positive value (PPV) of this classifier is 0.68 and negative predictive value (NPV) is 0.97.

**Conclusions:** Self-reported quantitative olfactory changes, either alone or combined with other symptoms, provide a specific and powerful tool for clinical diagnosis of COVID-19. The applicability of this tool for prioritizing COVID-19 laboratory testing is facilitated by a simple calculator presented here.

## BACKGROUND

In December 2019, the severe acute respiratory syndrome coronavirus 2 (SARS-CoV-2) was reported in Wuhan, China[1]. The resulting coronavirus disease COVID-19 has become a global pandemic with 16.5 million reported cases as of July 29th, 2020 (World Health Organization, 2020). When assessing SARS-CoV-2 infection, clinicians initially focused on the most common symptoms at the onset of COVID-19 illness such as fever, cough, and fatigue. Other reported signs and symptoms included sputum production, headache, hemoptysis, diarrhea, and dyspnea [2].

Since March 2020, an increasing number of reports regarding taste and smell loss in COVID-19 infections appeared in preprints[3, 4] and in general press, and it is currently well established that taste and smell loss is common in COVID-19 patients[5–8]. Earlier studies have already suggested associations between anosmia (loss of smell) and the coronavirus causing Severe Acute Respiratory Syndrome (SARS), SARS-CoV-1. Olfactory symptoms[9] and taste disorders[10] have also been associated also with viral upper respiratory tract infections caused by other viruses, as well. However, the prevalence of olfactory loss in COVID-19 is usually reported as much higher[11–15] than in other diseases[16]. In a recent crowd-sourced study, ∼7000 app users reported testing positive for COVID-19, with 65% of those reporting that they lost their sense of smell or taste[14], a three-fold increase in prevalence compared to COVID-19 negatives[17]. The severity of smell and taste loss in COVID-19 patients is striking: these sensory abilities were reduced by -79.7 ± 28.7, -69.0 ± 32.6 (mean ± SD), respectively, as reported by about 4000 participants using a 0-100 visual analog scale (VAS)[7].A follow-up study found that recent smell loss is the best predictor for COVID-19[18].

Here we assess the prevalence of different COVID-19 symptoms as well as the degree and additional characteristics of smell and taste changes in PCR-swab tested COVID-19 positive vs COVID-19 negative patients. Importantly, patients were recruited in a manner that did not disclose the underlying chemosensory questions in this study. We used these data to develop a classifier that can prioritize patients for PCR-testing, help epidemiological investigations, and screen large populations.

## METHODS

### Aim and setting

This prospective study compared symptoms in real time (RT) polymerase chain reaction (PCR)-tested COVID-19 positive and COVID-19 negative patients. Patients having PCR-test results (positive or negative) were recruited via social media (Twitter and Facebook) and word of mouth from March 2020 to May 2020 and interviewed from April 2020 to June 2020. The cohort comprised 224 Israeli patients aged ≥18 years (Figure 1) Israeli patients. The participants were not aware that the questionnaire will include smell and taste loss symptoms prior to their agreement to partake in the study. Informed consent was obtained from all participants. The study was approved by the Hadassah Medical Center Helsinki Committee (permit number 0236-20-HMO).

**Figure 1.**
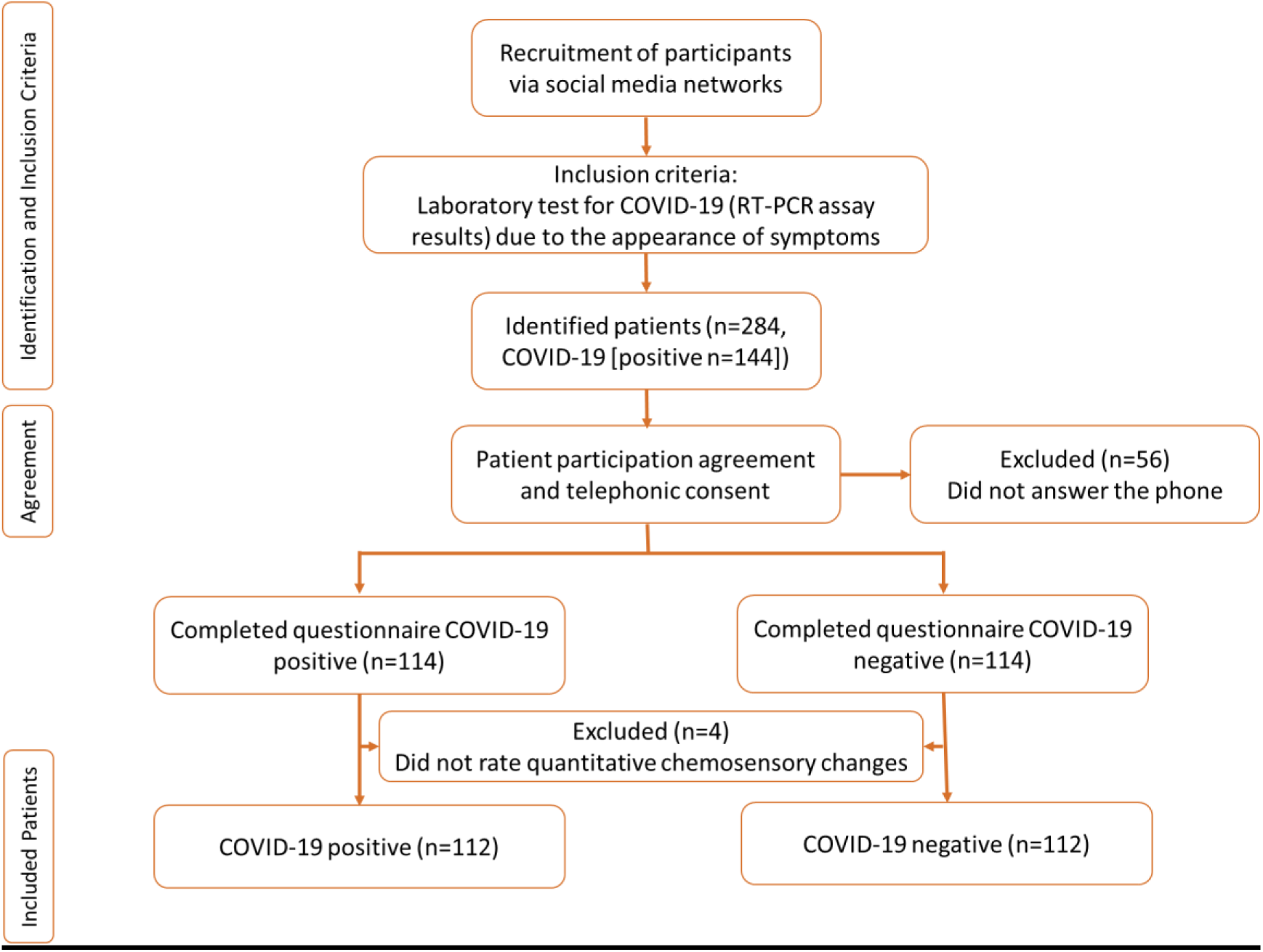
Flow chart of patients’ selection for case control study.

### General design

The interviews were carried out over the phone. The questionnaire is based on questions compiled by physicians and scientists in the Global Consortium for Chemosensory Research, GCCR[7]. The full questionnaire is included in Supplementary data and has five parts: 1) General information (e.g., age, gender); 2) Medical history (e.g., medical conditions, medications, changes in taste/smell in the past, pregnancy, contact with a confirmed patient); 3) Current illness: 23 physical signs and symptoms, including binary question (yes/no) on smell, taste and chemesthesis (cooling, burning, tingling sensation), PCR swab test results and dates, date of exposure to confirmed COVID-19 patient, subjective recovery feeling; 4) Smell: Participants were instructed to rate their sense of smell/ taste and the degree of their nose blockage on a scale from 1-10 (1 corresponding to “no sense of smell” and 10 to excellent sense of smell) and similarly rate the ability to breathe through the nose before/during/after illness. Blocked nose rating was used to test the plausible hypothesis that it causes a change of smell; 5) Taste (e.g., rating of taste ability before/during/after illness, as described for smell), experience of strange/bad taste in the mouth, change in sensitivity to irritants (chemesthesis) and change in basic taste modalities – sweet, salty, sour, bitter, each elicited by non-volatile compounds via specific receptors or channels expressed in dedicated taste receptor cells [19]. The fifth basic taste modality, umami or savory, was not used because it does not have a Hebrew translation. “Other” taste was available as an additional optional answer. Data was kept in Compusense Cloud on-line software (Compusense Inc., Guelph, ON, Canada).

### Statistical analysis

Log-odds for the individual symptoms were calculated over the full dataset. Confidence intervals and p-values for the log-odds were estimated from the *glm* function using the logistic link implemented in the statistical software R (https://www.r-project.org/).

Classifiers were trained from the reported symptoms to evaluate the separation between COVID-19 positive and COVID-19 negative patients. The classifiers were trained on a random subset of 2/3 of the data (the training set, 148 samples), and evaluated on the remaining samples (the test set). Sampling of the train and test sets was stratified by COVID-19 status. We trained the classifier on the full symptom matrix: All symptoms of question 23 in the questionnaire (see Supplementary) were included, except “no symptoms” or “other”. All eye symptoms were combined to “Eye symptoms”. Also added were quantitative questions for taste, smell, and nose blockage (rating before the illness minus rating during the illness, questions 31, 35, 37, 38, 40, 41 in the questionnaire) and chemesthesis (question 45) “Coated tongue”, “Dizziness”, “Ears pressure”, “Eye burn”, “Eye discharge”, “Hearing change”, “Lacrimation”, and “Vision changes” were removed since these symptoms were reported by less than 10% of the subjects. The classifiers were trained as penalized logistic regressions, using the elastic net algorithm (α = 0.5 implemented in the *glmnet* package in the R environment. This regression method encourages sparse coefficient vectors, meaning that it is suitable in situations where only few coefficients are non-zero. The regularization parameter (lambda) was initially set using cross-validation, but then increased until the model included no more than six symptoms. For classifiers based on a single symptom, no regularization was used.

Classifiers were evaluated using the hold-out test set. The score from the classifier was thresholded at zero, so that patients with score exceeding 0 were called positive by the classifier. Sensitivity (predicting COVID-19 positives correctly) and specificity (predicting COVID-19 negatives correctly) metrics were calculated from the following formulas:

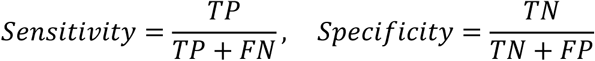

in which TP (TN) are the COVID-19 positives (negatives) classified correctly, and FN (FP) are the COVID-19 positives (negatives) classified incorrectly. Due to our balanced sample, accuracy is the average of sensitivity and specificity. We further computed the accuracy metrics that account for the expected proportion of positive cases in the tested population, namely the positive predictive value (PPV) and the negative predictive value (NPV). The scores obtained from the logistic classifier (*s*) were translated into probability to be positive (*P*) by adjusting for the proportion of COVID+ out of the tests (π) according to the following formulas:

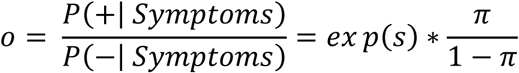

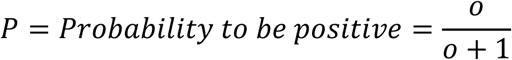

We take π to be 0.08 because that was the proportion during data collection.

The ROC curve corresponds to true-positive and false-positive rates for different values of the threshold; the curve and the area under the ROC curve (AUC), which measures the degree of separability between positive and negative scores, were estimated using the *pROC* package[20].

### Sample size calculation

The sample size was calculated to allow detecting differences in smell-loss or taste-loss prevalence between COVID-19 positive and negative populations. Based on previous research [e.g. [21]], we used conservative estimates of 60% prevalence in the positive population and 35% prevalence in the negative population. Power was estimated by Monte Carlo simulations, namely repeatedly (b=1000) resampling from two Binomial distributions corresponding to the positive population and the negative population. Assuming 100 individuals are assigned to each group, and a two-sided t-test is used, the probability of detection (power) is 92%. To be on the conservative side, we used somewhat larger samples.

## RESULTS

### Patients’ characteristics

Completed questionnaires were obtained from 112 COVID-19 positives and 112 COVID-19 negatives. The median age of the respondents was 35 ± 12 years for positives and 37 ± 12 years for negatives (mean ± SD) years. The positives group included more men (64%), while the negatives group was more balanced (48% males). Seven patients classified as hospitalized (received respiratory support during their hospitalization and / or were hospitalized in the intensive care unit) and the rest 217 were classified as ambulatory patients.

### Patients’ symptoms

Signs and symptoms that appeared in the binary part of the questionnaire (Supplementary Material, question 23 of the full questionnaire) and were found to occur in at least 10% of the positive patients are summarized in Table 1. A few symptoms, including dry cough and sore throat, were prevalent in COVID-19 positives, but even more so in the negatives control sample.

**Table 1.**
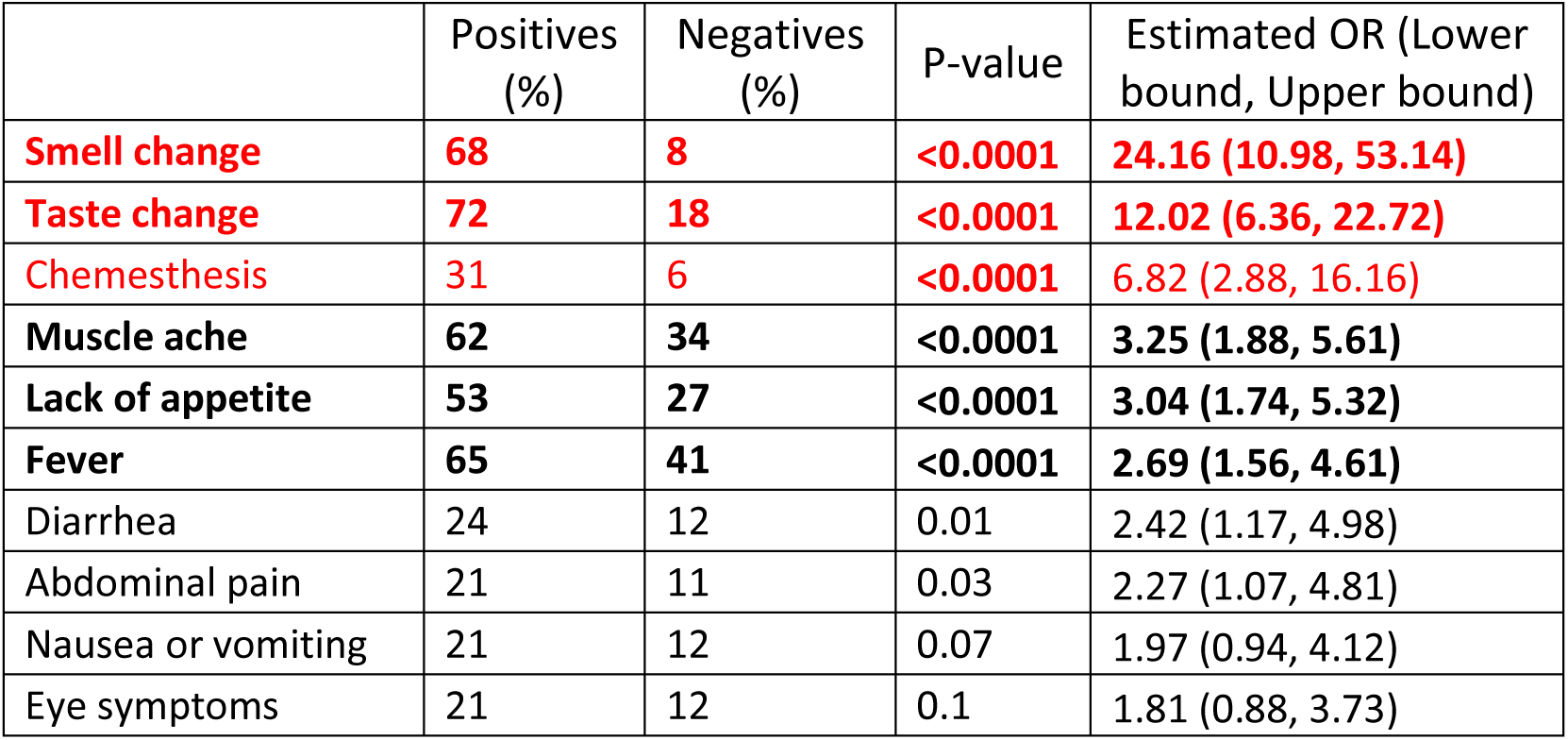
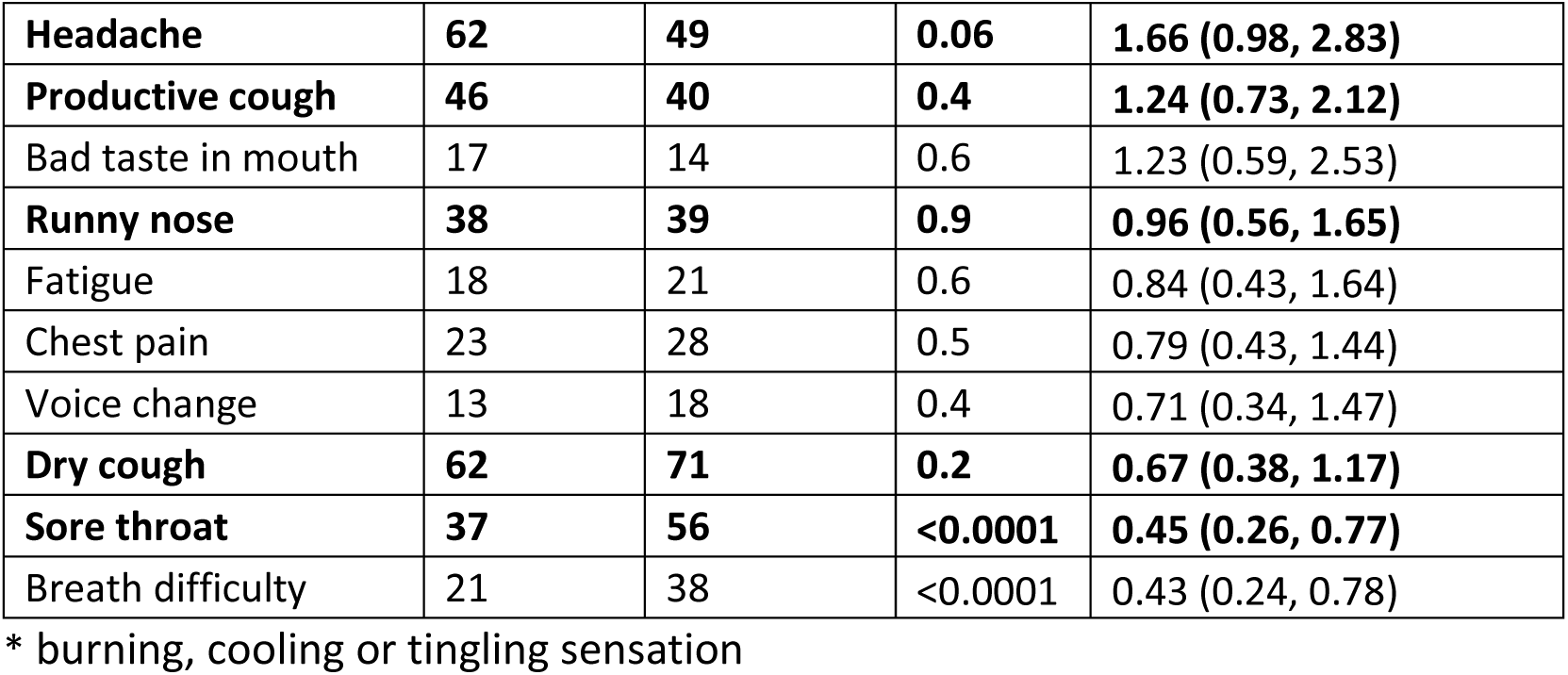
Prevalence and odds ratios of signs and symptoms. Only signs and symptoms that were present in at least 10% of all study participants are listed. Those with the highest odds ratios are shown in red and those reported by at least a third of positives are shown in boldface.

Smell change, taste change, change in chemesthetic ability (perceiving spicy, tingling or cooling sensations) and muscle ache were significantly more prevalent in COVID-19 positive as compared to COVID-19 negative patients (68%, 72%, 31%, 62% vs. 8.0%, 18%, 6%, 34%, respectively) (Table 1). Other CDC recognized symptoms[22], such as lack of appetite, fever, and diarrhea were approximately twice or three times more common among positives than negatives.

Nausea or vomiting, although considered a COVID-19 symptom, were not more common among COVID-19 positive as compared to COVID-19 negative patients. By contrast, lack of appetite, despite not being included as an “official” CDC symptom[22], was found to be significantly more common in COVID-19 positive patients.

Taste and smell change often, but not always, together: Figure 2A shows the distribution of reports on taste and smell changes. Change in both smell and taste perception was reported by 63% of positive patients and only 6% of negative patients. 4% of positive patients experienced only smell change and 9% reported taste change with no smell change.

**Figure 2.**
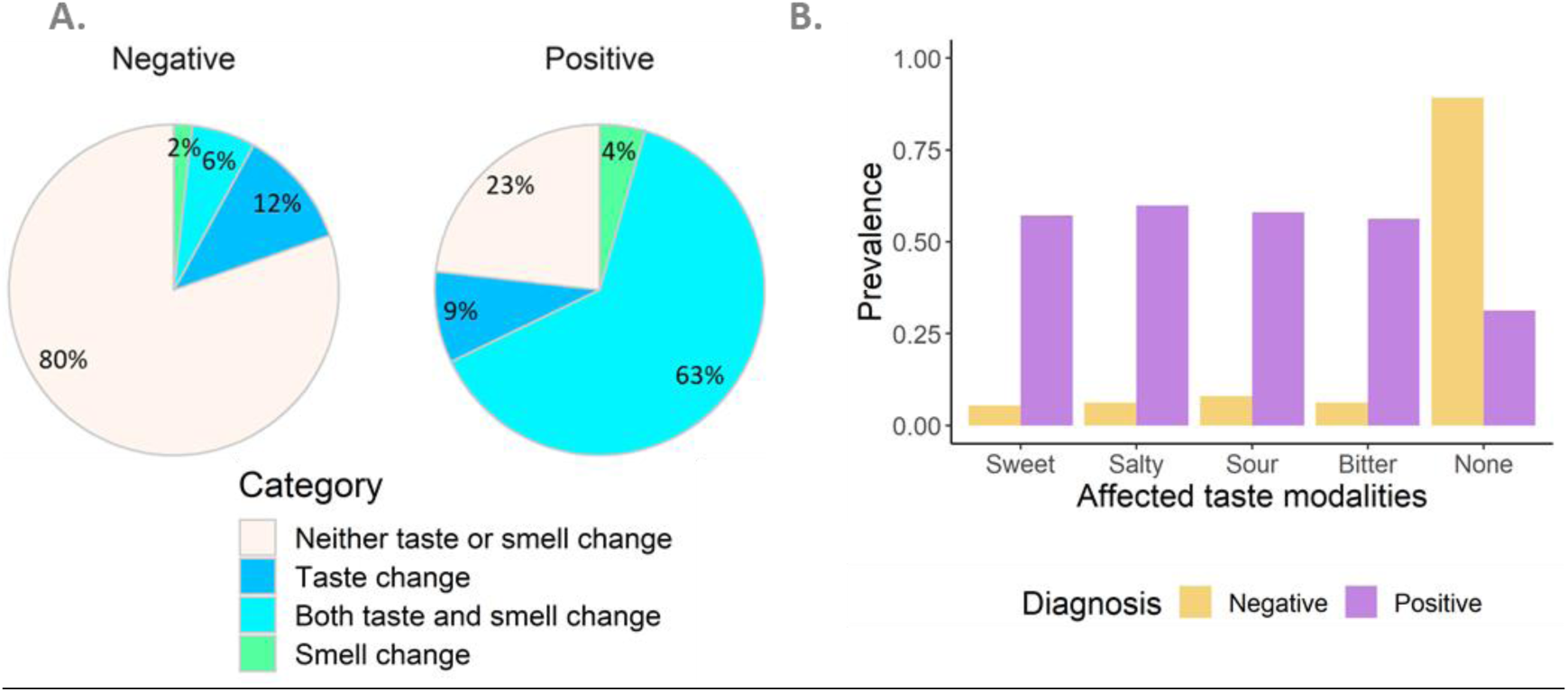
Smell, taste, and taste modalities changes during disease. **A**. The pie chart presents prevalence of smell and taste changes in positive and negative patients, occurring together or separately. Numbers indicate percentage of COVID-19 positives and negatives reporting taste or smell loss (blue and green respectively), both taste and smell loss (turquoise) or neither (seashell). **B**. The prevalence of the four taste modalities in COVID-19 positive and negative patients. COVID-19 positives are represented in purple and COVID-19 negatives are represented in orange

Approximately 60% of the positive patients reported impairment of at least one of the four taste modalities (sweet, salty, sour, and bitter) compared to only ∼6% among the negative patients (Figure 2B). In COVID-19 positive patients with taste impairment, all four taste modalities were usually impaired. In 31% of the positive patients and in 90% of the negative ones, no taste modality was impaired.

### Chemosensory changes rating and details

In addition to the binary questions, the participants were asked to rate their smell and taste senses before and during their illness on a 1-10 scale.

As is seen in Figure 3, the change in smell and taste ability during disease, compared to a self-reported individual baseline before the disease, differs greatly between positive and negative patients. Both taste and smell changes were significantly greater for COVID-19 positive patients compared to COVID-19 negative patients (4.5±3.6 and 4.9±3.8 vs. 0.5±1.5 and 0.4±1.5, mean ± SD, p<0.0001 and p<0.0001, respectively). When considering only patients who reported taste or smell changes (answered “yes” to the respective binary questions), the averages in positive patients were 6.3±2.6 for taste and 7.1±2.4 for smell, compared to negative patients with 2.1±2.3 for taste and 4.8±2.6 for smell (among patients with any change in taste p<0.0001, in smell p<0.0001).

**Figure 3.**
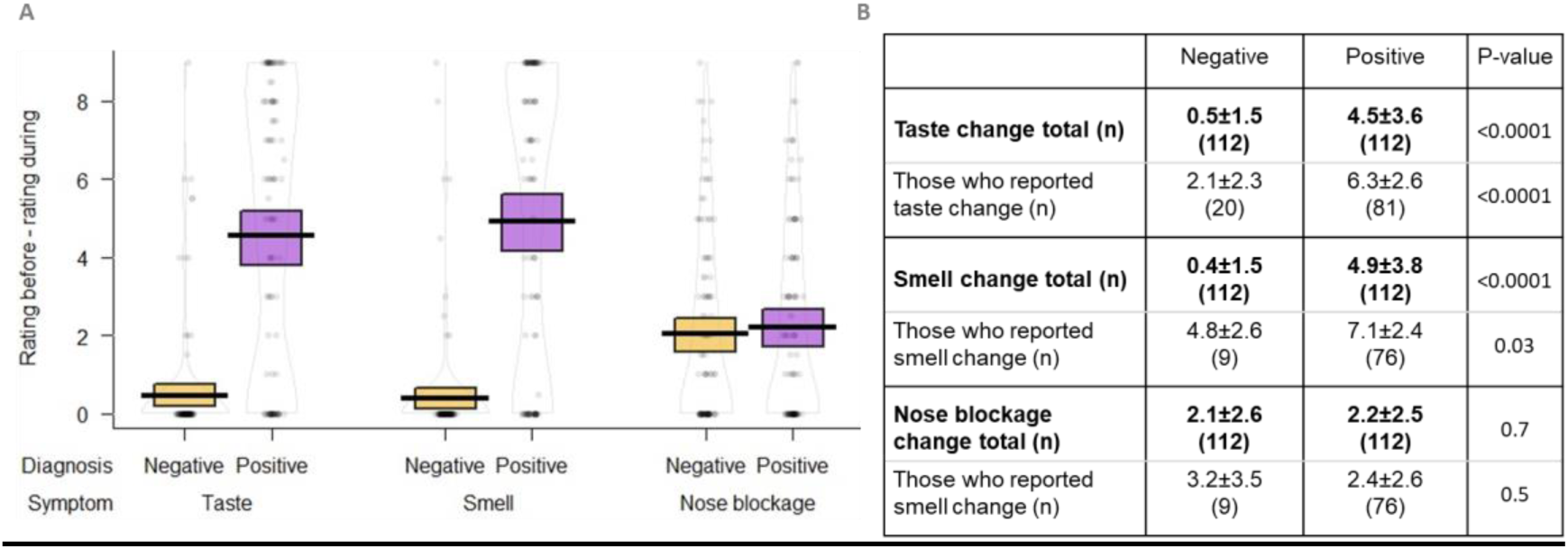
Degree of taste smell and nose blockage in COVID-19 positives and negatives. **A**. Histogram showing the change in rating during the illness minus rating before the illness for taste, smell, and nose blockage. No change is coded 0 and the highest change is 9. COVID-19 positives are represented in purple and COVID-19 negatives are represented in orange. **B**. Table of mean ± SD for COVID-19 positives and negatives in general and for those reporting changes of taste or smell. Scores for taste, smell, and nose blockage were evaluated on a 1-10 scale. P values for the difference in the magnitude of change between COVID-19 positives and negatives was calculated using a two-sided t-test.

We next set out to check which combination of symptoms will be most useful for differentiating between COVID-19 positive and COVID-19 negative diagnosis. To that end, several classifiers were trained based on 66% of the sample and evaluated on 34% that was kept as a holdout set. The process of selection of descriptors is outlined in Figure 4. Relevant symptoms (n=30) were included as possible descriptors for the classifiers, and the elastic-net penalization was increased until no more than six symptoms were included in the model (number limited for practicality). The effect of excluding or including a particular symptom was evaluated in order to understand the importance of separating taste from smell and using binary vs. quantitative measures for each (Figure 4).

**Figure 4.**
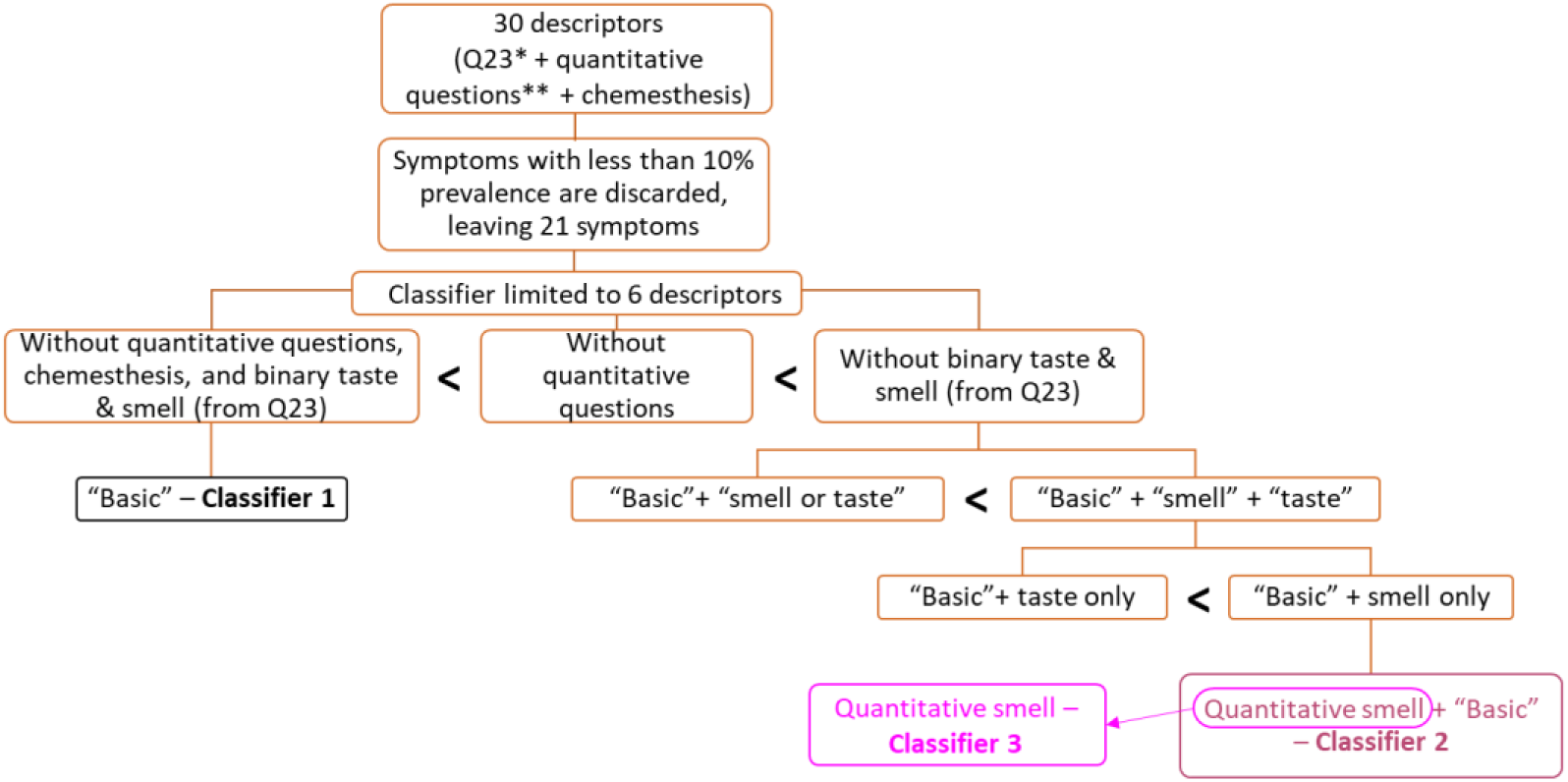
Descriptors selection process for classifiers. The flow chart depicts total symptoms selected for the classifiers. Upon limiting the number of descriptors for the classifiers and excluding chemosensory symptoms as described, Classifier 1 was created. Different combinations of symptoms established better classifiers than Classifier 1, those using quantitative questions exhibiting better performance than those using binary ones. The classifier using “smell” **and** “taste” as separate descriptors, rather than “smell **or** taste” as a single joint descriptor showed better performance. The “Basic” + smell only descriptor outperformed the “Basic” + taste only descriptor, resulting in Classifier 2. Finally, the smell only descriptor was tested alone without all other “Basic” symptoms, resulting in Classifier 3.

The results of the evaluation on the holdout set are summarized in Supplementary Table 1, and classifiers 1-3 can be seen in Figure 5. Classifiers that did not use chemosensory symptoms had poor performance (AUC 0.60, black curve, classifier 1, and additional classifiers (Table S1). Adding the quantitative smell-change symptom (maroon curve, Classifier 1) is sufficient to outperform all other classifiers (AUC 0.83). Remarkably, using quantitative smell-change as a sole symptom (magenta curve, Classifier 3) resulted in a classifier that was nearly equally effective as Classifier 2 (AUC 0.81).

**Figure 5.**
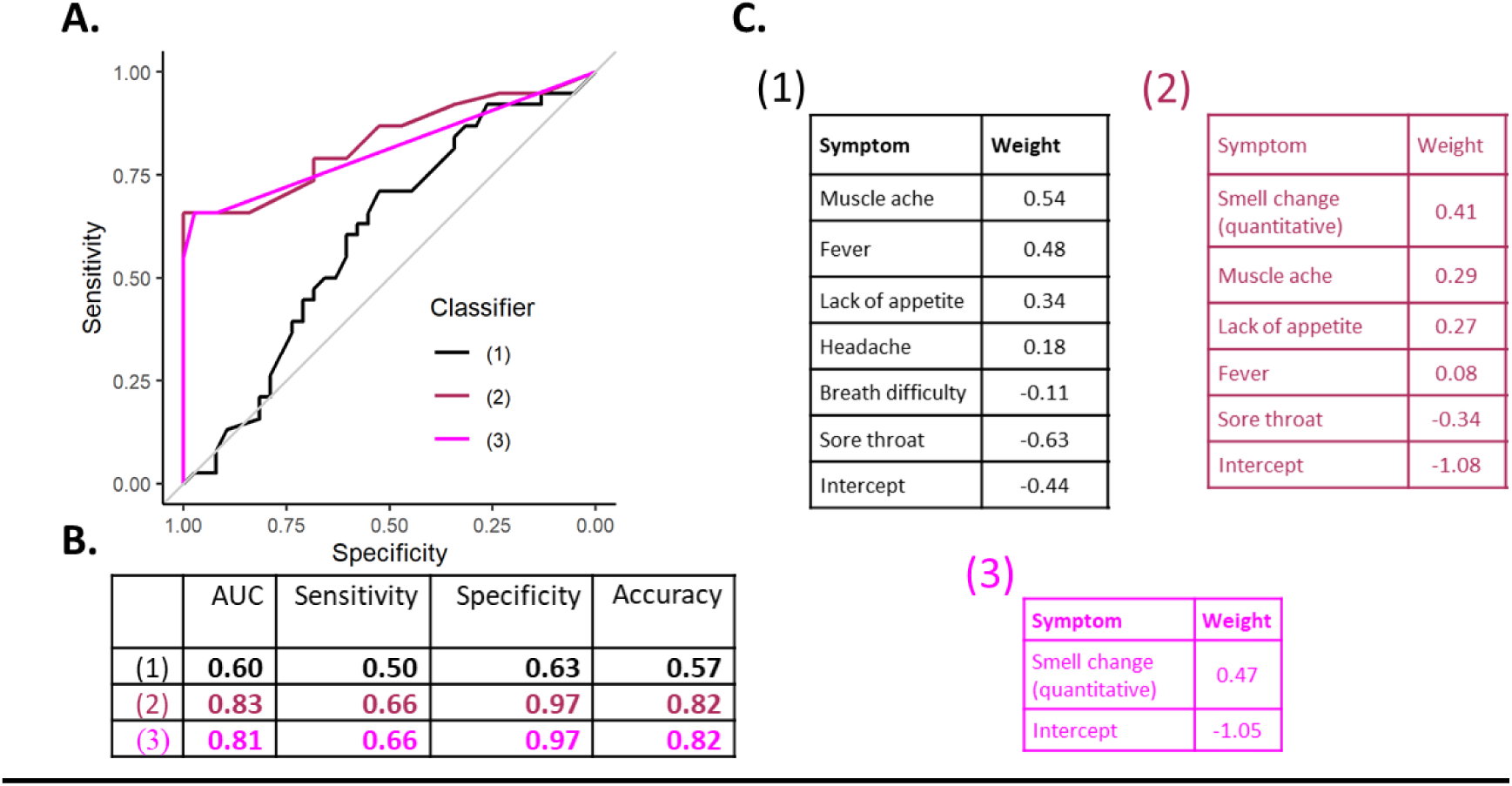
**A**. ROC curves for the classifiers considered. **B**. Statistical parameters for the ROC curves. For the quantitative descriptors, the coefficients correspond to a descriptor scaled to the 0-1 range. **C**. Descriptors, their coefficients and intercept (fitted parameter) used in Classifiers 1, 2 and 3.

Adding taste change to Classifier 2 did not improve its performance, as it resulted in AUC of 0.82 (Classifier 7, Table S1). Furthermore, taste change as a sole descriptor resulted in AUC of 0.75 (Classifier 15, Table S1) and as an added descriptor to other “Basic” symptoms, it resulted in AUC of 0.76 (Classifier 13, Table S1). Thus, while there is a high correlation (0.82) between quantitative changes in smell and quantitative changes in taste, in our sample the smell change descriptor outperforms the taste descriptor. Using the quantitative smell and taste descriptors resulted in higher AUC’s than binary (yes/no) descriptors of these changes. For example, a binary smell descriptor used as a sole descriptor resulted in AUC of 0.78 (Classifier 16, Table S1), as compared to AUC of 0.81 using quantitative smell descriptor.

Scores according to the classifiers can be readily transformed into the probability for COVID-19, under additional factor: the known rate of positive tests. The rate of positive tests out of total PCR tests at the time of participants recruitment was calculated to be 8% based on data from Israeli Ministry of Health. By incorporating this factor (as described in the Methods), we were able to reach PPV of 0.68 and NPV of 0.97 for both classifiers. A probability calculator based on Classifiers 2, 3 and 13 (from Supplementary) is available in GITHUB (https://github.com/KimAsseo/Hadassah_COVID-19) and may be used for practical purposes in clinical life.

## DISCUSSION

We have established the prevalence and degree of decrease in taste and smell in patients who were eligible to receive PCR swab tests during the COVID-19 pandemic and found significant differences in PCR-positive and PCR-negative patients.

The change in smell ability is not related to nasal obstruction, as nose blockage was low, in accord with previous studies[7, 23]. Taste and chemesthesis changes strongly correlate with smell change (also shown and discussed by Parma[7]). Taste changes are more common in negatives, and chemesthesis are less common in positives, leading to odds ratios for these chemosensory modalities that are high, but lower than for olfaction. Accordingly, taste and chemesthesis were not captured by the classification analysis as characteristics contributing to improvement of the classifier. All taste modalities in COVID-19 patients were impacted together (or not at all). This is of interest for understanding the pathophysiology of the disease: a recent study suggests CoV-2 infection of non-neuronal cell types expressing ACE2 and TMPRSS2 as the mechanism underlying COVID-19 related anosmia[24], but the reason for COVID-19 ageusia is less clear [19, 25]. Our results support the idea of impairment of supporting cells or tissues, rather than of taste receptors cells Type 2, which express bitter, sweet and umami taste receptors or Type 3 which express sour sensing channels [26]. However, our observation that individual taste modalities and general taste ability are greatly impaired in COVID-19 patients did not warrant addition of taste questions for patients screening, as these did not contribute to the classifier performance. Nevertheless, patients with prior conditions of impaired olfaction (estimated 5% of population[27]) require a suited classifier.

We thus present three versions of classifiers: the first one is based on four yes/no questions (muscle ache, lack of appetite, fever, sore throat) and the quantitative smell change calculated from self-rating of smell perception before and during the current illness. The second version uses only the quantitative smell change and has a similar AUC. The third one (suited for participants with pre-existing olfactory impairment) is based on five yes/no questions (muscle ache, lack of appetite, fever, sore throat, breath difficulty) and quantitative taste change. The probabilities for COVID-19 based on these calculators are available via Github.

Our study confirms previous reports [21, 28] showing an association between changes in smell and taste with the positive status of COVID-19. Our best-preforming classifier (Classifier 2, AUC 0.83) used symptoms found to be predictive also by Menni *et al*.[14]. Specifically, these authors reported chemosensory loss and loss of appetite as positively contributing. However, their chemosensory descriptor is “loss of smell and taste”, while ours involves quantitative smell change.

Our study was performed in parallel to Gerkin *et al*.[18], and similarly included both binary and quantitative questions regarding taste and smell as two separate indicators. Our results for positive patients are in overall agreement with Parma *et al*.[7] and Gerkin *et al*.[18]. This is striking in view of the different methods employed for recruitment (targeting smell and change impairment worldwide vs PCR-positive COVID-19 patients in Israel), data collection (online survey vs telephone interviews), and quantitative scales (100-point visual analogue scales (VAS) vs. 1-10 scale in this 0/82study). In essence, both Gerkin *et al*. and the current study suggest that quantitative smell change is the best predictor of COVID-19 in single and in cumulative feature models and is better than binary feature. The superior performance of the classifier (AUC of 0.83 vs. 0.72 in [18]) is probably due to a more realistic representation of our sample, in which chemosensory losses were not over-represented.

With the increasing public awareness to smell impairment as COVID-19 characteristic symptoms, individuals presenting smell and taste change are now more likely to be suspected of having COVID-19. By considering other symptoms and the severity of chemosensory change, our calculators provide a free, fast and easy to use tool that can provide immediate answers for patients awaiting their PCR-swab test results and potentially decrease anxiety of negative patients who experienced smell and taste impairments.

### Study limitations

The method of patient recruitment is one of the limitations of this study: social media-based recruitment may limit participants’ representation as it targets mostly younger patients, with internet access and social media accounts. Word of mouth recruitment was used as well and contributes as well to creating a sample that is not necessarily representative of the general population. Male and female patients were not fully matched across positives (64% males) and negatives (48% males), in accord with higher % of males (56%) among COVID-19 patients in Israel.

Importantly, symptoms-based classifiers cannot capture asymptomatic COVID-19 patients. Therefore, low probability established with our classifiers should not be considered as a predictor of negative COVID-19 status. In other words, our classifiers are not SNOUT (‘Sensitive test when Negative rules OUT the disease’) but can definitely be referred to as ‘Specific test when Positive rules IN the disease’ (SPIN).

While chemosensory loss is a dominant feature of symptomatic COVID-19 patients, about 30% of them do not report such loss. Our sample was not large enough to include many positive patients in this subgroup, and further studies are needed to capture distinctive characteristics of COVID-19 patients with intact smell and taste.

Additionally, our sample was composed of light to moderately ill patients, thus the classifiers reported are not necessarily applicable to patients with severe forms of COVID-19. It should also be kept in mind that our data is specific to Israeli patients and reflect to some degree the criteria for PCR tests eligibility during the recruitment (fever and dry cough were sufficient for PCR test but change of smell and/or taste alone was not).

Another possible limitation of this study is the self-reporting, rather than objective testing method for data collection used. Clearly, the infectious nature of the disease cause data collection to be mostly based on self-reports[29], but classifiers based on brief objective examination of taste and smell symptoms may provide additional insights.

Lastly, RT-PCR is a commonly used diagnostic test for COVID-19 and for that reason it was used in the present study as a distinguishing criterion. Nevertheless, even this test may report false diagnosis (26%-37%)[30, 31]. Considering the inaccuracy of PCR tests, high scores from our classifiers may help to reinforce true positive results and possibly capture PCR false negative patients.

## CONCLUSIONS

The resurging pandemic puts the clinic and public health authorities in a scenario not usual for modern medicine – namely, the limited resources require or may require in the future, prioritization of testing and treatment. The fact that our sample contained PCR-positive and PCR-negative ambulatory patients, all suspected to have COVID-19 prior to PCR testing, enabled the development of symptoms-based classifiers.

Our results suggest that ranking of the ability to smell before and during the illness, is an excellent practical approach to identify COVID-19 positive patients offering reasonably high predictive capability (Specificity 97%, Accuracy 82%). Additional classifier is available for patients with prior olfactory impairments (Supplementary Material Table S2).

Based on the classifiers developed in this work, we propose a simple calculator that can be used to prioritize testing (available at https://github.com/KimAsseo/Hadassah_COVID-19). Additionally, high-performance classifier may potentially capture false negative PCR tests results of high scored individuals. Detection and isolation of potential COVID-19 patients is of crucial importance for pandemic containment but remains a persistent challenge worldwide due to lacking accessibility to tests. While further research is required, the current study provides a practical tool for assessing potential COVID-19 patients.

## Data Availability

COVID-19 probability calculators based on developed classifiers are available at GITHUB

https://github.com/KimAsseo/Hadassah_COVID-19

## LIST OF ABBREVIATIONS

CDC: Centers for Disease Control and Prevention
SARS-CoV-2: Severe Acute Respiratory Syndrome Coronavirus 2
SARS: Severe Acute Respiratory Syndrome
VAS: Visual Analog Scale
SD: Standard Deviation
GCCR: Global Consortium for Chemosensory Research
AUC: Area Under the Curve
PPV: Positive Predictive Value
NPV: Negative Predictive Value
RT: Real Time
PCR: Polymerase Chain Reaction
SNOUT: Sensitive test when Negative rules OUT the disease
SPIN: Specific test when Positive rules IN the disease
ROC curve: Receiver Operating Characteristic curve

## AUTHORSHIP CONTRIBUTION

NK and MYN initiated the research, NK and MN recruited patients, NK, MYN, YB and MM and designed the research, SI, MM, RNP and MYN supervised the research, HK carried out the interviews, HK and MYN drafted the manuscript, KA and YB preformed the statistical analysis and created figures and tables. All authors contributed to writing and approved the final manuscript.

## ACKNOWLEDGEMENTS

MYN is supported by Masha Niv is supported by Israel Science Foundation grant #1129/19. HK is a recipient of the Uri Zehavi Scholarship.

## Ethics approval and consent to participate

The study was conducted in accordance with Helsinki committee and the required ethics approval was granted (reference number HMO-0236-20).

## Consent for publication

Written informed consents for publication of patients’ clinical details were obtained from the patients.

## BIBILIOGRAPHY

1. Zhu N, Zhang D, Wang W, Li X, Yang B, Song J, et al. A novel coronavirus from patients with pneumonia in China, 2019. N Engl J Med. 2020;382:727–33. doi:10.1056/NEJMoa2001017.

2. Rothan HA, Byrareddy SN. The epidemiology and pathogenesis of coronavirus disease (COVID-19) outbreak. Journal of Autoimmunity. 2020;109:102433.

3. Machado C, Gutierrez JV. ANOSMIA AND AGEUSIA AS INITIAL OR UNIQUE SYMPTOMS AFTER SARS-CoV-2 VIRUS INFECTION (Review article). 2020;2 April. doi:10.20944/preprints202004.0272.v1.

4. Levinson R, Elbaz M, Ben-Ami R, Shasha D, Levinson T, Choshen G, et al. Title: Anosmia and dysgeusia in patients with mild SARS-CoV-2 infection. Cold Spring Harbor Laboratory Press; 2020. doi:10.1101/2020.04.11.20055483.

5. Moein ST, Hashemian SMR, Mansourafshar B, Khorram-Tousi A, Tabarsi P, Doty RL. Smell dysfunction: a biomarker for COVID-19. Int Forum Allergy Rhinol. 2020;:alr.22587. doi:10.1002/alr.22587.

6. Gilani S, Roditi R, Naraghi M. COVID-19 and anosmia in Tehran, Iran. Med Hypotheses. 2020;141:109757.

7. Parma V, Ohla K, Veldhuizen MG, Niv MY, Kelly CE, Bakke AJ, et al. More than smell – COVID-19 is associated with severe impairment of smell, taste, and chemesthesis. Chemical Senses | Oxford Academic. 2020.https://academic.oup.com/chemse/advance-article/doi/10.1093/chemse/bjaa041/5860460. Accessed 22 Jun 2020.

8. Whitcroft KL, Hummel T. Olfactory Dysfunction in COVID-19: Diagnosis and Management. JAMA - Journal of the American Medical Association. 2020.

9. Soler ZM, Patel ZM, Turner JH, Holbrook EH. A primer on viral-associated olfactory loss in the era of COVID-19. International Forum of Allergy and Rhinology. 2020.

10. Bay E. Smell and taste disorders. Med Welt. 1961;42:2143–7.

11. Bagheri SHR, Asghari AM, Farhadi M, Shamshiri AR, Kabir A, Kamrava SK, et al. Coincidence of COVID-19 epidemic and olfactory dysfunction outbreak. medRxiv. 2020;:2020.03.23.20041889.

12. Lechien JR, Chiesa-Estomba CM, De Siati DR, Horoi M, Le Bon SD, Rodriguez A, et al. Olfactory and gustatory dysfunctions as a clinical presentation of mild-to-moderate forms of the coronavirus disease (COVID-19): a multicenter European study. Eur Arch Oto-Rhino-Laryngology. 2020.

13. Mao L, Jin H, Wang M, et al. Neurologic Manifestations of Hospitalized Patients With Coronavirus Disease 2019 in Wuhan, China [published online ahead of print, 2020 Apr 10]. JAMA Neurol. 2020;77(6):1–9. doi:10.1001/jamaneurol.2020.1127

14. Menni C, Valdes AM, Freidin MB, et al. Real-time tracking of self-reported symptoms to predict potential COVID-19. Nat Med. 2020;26(7):1037–1040. doi:10.1038/s41591-020-0916-215.

15. Yan CH, Faraji F, Prajapati DP, Boone CE, DeConde AS. Association of chemosensory dysfunction and COVID-19 in patients presenting with influenza-like symptoms. Int Forum Allergy Rhinol. 2020;10(7):806–813. doi:10.1002/alr.22579

16. Tong JY, Wong A, Zhu D, Fastenberg JH, Tham T. The Prevalence of Olfactory and Gustatory Dysfunction in COVID-19 Patients: A Systematic Review and Meta-analysis. Otolaryngol Neck Surg. 2020;:019459982092647. doi:10.1177/0194599820926473.

17. Menni C, Sudre CH, Steves CJ, Ourselin S, Spector TD. Quantifying additional COVID-19 symptoms will save lives. Lancet (London, England). 2020;0. doi:10.1016/S0140-6736(20)31281-2.

18. Gerkin RC, Ohla K, Veldhuizen MG, Joseph P V., Kelly CE, Bakke AJ, et al. Recent smell loss is the best predictor of COVID-19: a preregistered, cross-sectional study. medRxiv. 2020.

19. Chaudhari N, Roper SD. The cell biology of taste. Journal of Cell Biology. 2010.

20. Robin X, Turck N, Hainard A, Tiberti N, Lisacek F, Sanchez JC, et al. pROC: An open-source package for R and S+ to analyze and compare ROC curves. BMC Bioinformatics. 2011;12:77. doi:10.1186/1471-2105-12-77.

21. Menni C, Valdes A, Freydin MB, Ganesh S, Moustafa JE-S, Visconti A, et al. Loss of smell and taste in combination with other symptoms is a strong predictor of COVID-19 infection. medRxiv. 2020;:2020.04.05.20048421.

22. Symptoms of Coronavirus | CDC. https://www.cdc.gov/coronavirus/2019-ncov/symptoms-testing/symptoms.html. Accessed 24 Jul 2020.

23. Salmon D, Bartier S, Hautefort C, Nguyen Y, Nevoux J, Hamel A-L, et al. Self-reported loss of smell without nasal obstruction to identify COVID-19. The multicenter CORANOSMIA cohort study. J Infect. 2020. doi:10.1016/j.jinf.2020.07.005.

24. Brann DH, Tsukahara T, Weinreb C, Lipovsek M, Van den Berge K, Gong B, et al. Non-neuronal expression of SARS-CoV-2 entry genes in the olfactory system suggests mechanisms underlying COVID-19-associated anosmia. Sci Adv. 2020;:eabc5801. doi:10.1126/sciadv.abc5801.

25. Cooper KW, Brann DH, Farruggia MC, Bhutani S, Pellegrino R, Tsukahara T, et al. COVID-19 and the chemical senses: supporting players take center stage. Neuron. 2020;0. doi:10.1016/j.neuron.2020.06.032.

26. Roper SD, Chaudhari N. Taste buds: Cells, signals and synapses. Nature Reviews Neuroscience. 2017;18:485–97. doi:10.1038/nrn.2017.68.

27. Hummel T, Landis BN, Hüttenbrink K-B. Smell and taste disorders. www.awmf.org/leitlinien/detail/ll/017-050.html. Accessed 19 Jul 2020.

28. Roland LT, Gurrola JG, Loftus PA, Cheung SW, Chang JL. Smell and taste symptom-based predictive model for COVID-19 diagnosis. Int Forum Allergy Rhinol. 2020;10:832–8. doi:10.1002/alr.22602.

29. Agyeman AA, Chin LL, Landersdorfer KB, Liew CB, Ofori-Asenso D. Smell and Taste Dysfunction in Patients With COVID-19: A Systematic Review and Metaanalysis. Mayo Clin Proc. 2020. doi:10.1016/j.mayocp.2020.05.030.

30. Sethuraman N, Jeremiah SS, Ryo A. Interpreting Diagnostic Tests for SARS-CoV-2. JAMA - Journal of the American Medical Association. 2020;323:2249–51.

31. Wang W, Xu Y, Gao R, Lu R, Han K, Wu G, et al. Detection of SARS-CoV-2 in Different Types of Clinical Specimens. JAMA - Journal of the American Medical Association. 2020;323:1843–4.

